# The implementation of remote home monitoring models during the COVID-19 pandemic in England

**DOI:** 10.1101/2020.11.12.20230318

**Authors:** Cecilia Vindrola-Padros, Manbinder S Sidhu, Theo Georghiou, Chris Sherlaw-Johnson, Kelly E Singh, Sonila M Tomini, Jo Ellins, Steve Morris, Naomi J Fulop

**Affiliations:** Department of Targeted Intervention, University College London (UCL), UK; Health Services Management Centre, School of Social Policy, University of Birmingham, Park House, UK; Nuffield Trust, London, UK; Department of Applied Health Research, University College London (UCL), London, UK; University of Cambridge, Cambridge, UK

## Abstract

**Background:** There is a paucity of evidence for the implementation of remote home monitoring for COVID-19 infection. The aims of this study were to identify the key characteristics of remote home monitoring models for COVID-19 infection, explore the experiences of staff implementing these models, understand the use of data for monitoring progress against outcomes, and document variability in staffing and resource allocation.

**Methods:** This was a multi-site mixed methods study that combined qualitative and quantitative approaches to analyse the implementation and impact of remote home monitoring models during the first wave of the COVID-19 pandemic (July to September 2020) in England. The study combined interviews (n=22) with staff delivering these models across eight sites in England with the collection and analysis of data on staffing models and resource allocation.

**Findings:** The models varied in relation to the healthcare settings and mechanisms used for patient triage, monitoring and escalation. Implementation was embedded in existing staff workloads and budgets. Good communication within clinical teams, culturally-appropriate information for patients/carers and the combination of multiple approaches for patient monitoring (app and paper-based) were considered facilitators in implementation. The mean cost per monitored patient varied from £400 to £553, depending on the model.

**Interpretation:** It is necessary to provide the means for evaluating the effectiveness of these models, for example, by establishing comparator data. Future research should also focus on the sustainability of the models and patient experience (considering the extent to which some of the models exacerbate existing inequalities in access to care).

**Funding:** The study was funded by the National Institute for Health Research-NIHR (Health Services and Delivery Research, 16/138/17 – Rapid Service Evaluation Research Team; or The Birmingham, RAND and Cambridge Evaluation (BRACE) Centre Team (HSDR16/138/31).

## INTRODUCTION

Delays in the presentation of patients with COVID-19 has led to patients arriving in acute care emergency departments with very low oxygen saturations, often without accompanying breathlessness (‘silent hypoxia’)^1^. As a result, many patients have been admitted with an advanced course of the disease, requiring extended hospital length of stays, invasive treatment, potential admission to Intensive Care Units (ICU) and death^2^. Remote home monitoring models (sometimes referred to as ‘virtual wards’), using pulse oximetery and other measurements (i.e. temperature), seek to remotely monitor patients considered at high-risk of deterioration at home to: 1) avoid unnecessary hospital admissions and promote appropriate admissions (appropriate care at the appropriate place), and 2) escalate cases of deterioration at an earlier stage to avoid invasive ventilation and ICU admission (by referring patients to emergency services or asking them to visit primary care)^3^. Remote home monitoring models have been implemented for confirmed or suspected COVID-19 cases in the US, Australia, Canada, Ireland, China, The Netherlands, India and the UK, with some variation in the frequency of patient monitoring, modality (telephone or video calls and use of applications or online portals), staffing, patient criteria and use of pulse oximetry to measure oxygen saturation levels^4-8^.

In England, a number of remote home monitoring models were set-up during the first wave of the COVID-19 pandemic with the aim outlined above but with a high degree of variability in relation to the mechanisms implemented for patient assessment and triage, monitoring and escalation^9^. Some models have been led by secondary care while others are mainly based in primary care. Furthermore, some have been designed as pre-hospital models (admitting patients from the community or emergency department) while others have functioned as an early discharge service from the hospital ‘wards’^9 10^.

Despite previous research on the use of remote home monitoring models for other conditions and their widespread use during the COVID-19 pandemic, studies on their implementation and impact are rare^11^. This mixed-methods evaluation of remote home monitoring models in England sought to address this gap by capturing the processes of implementing remote home monitoring models for confirmed or suspected COVID-19 cases, variability in staffing and resource allocation and the experiences of staff designing and delivering these models. The findings from this study were used to inform the planning for wave 2 of the pandemic (including the implementation of additional pilot sites), the national rollout of the Covid Oximetry@ Home programme by NHS England and Improvement and the design of an evaluation of this national programme.

The aims of this study were to: identify the key characteristics of remote home monitoring models, explore the experiences of staff implementing these models, understand the use of data for monitoring progress against outcomes, and document variability in staffing and resource allocation. We studied models with the following characteristics:implemented to monitor confirmed or suspected COVID-19 cases (retrospective as we focused on sites implemented during the first wave of the pandemic), focused on monitoring patients prior to hospital admission as well as early discharge from hospital wards, delivered from primary and secondary care settings and include some element of patient recording of oxygen saturation using pulse oximetry.

The research questions were:

1. What were the aims and main designs of the remote home monitoring models implemented for confirmed or suspected COVID-19 cases?
2. What were the factors that acted as barriers and facilitators in the implementation of these models during wave 1 of the pandemic?
3. What were the expected outcomes of these models?
4. What data were collected by study sites and how did these help them monitor progress against their expected outcomes?
5. How were the resources allocated (including staffing models) to implement the remote home monitoring models during wave 1 of the pandemic?
6. What were the lessons learned from implementing remote home monitoring models during wave 1 of the pandemic?

## METHODS

This was a rapid multi-site study that combined qualitative and quantitative approaches to analyse the implementation and impact of remote home monitoring models implemented for confirmed or suspected COVID-19 cases. The study was carried out from July to September 2020. The full study protocol can be accessed: https://www.nuffieldtrust.org.uk/rset-the-rapid-service-evaluation-team#2.

### Rapid qualitative study of first wave

Qualitative fieldwork was based on telephone and online semi-structured interviews with a purposive sample of staff from eight pilot sites implemented during the first wave of the pandemic and documentary analysis of internal documents developed by these sites.The purposive sampling strategy is described below. The selection of the eight pilot sites included in the study was also shaped by convenience sampling, as we identified sites through a Community of Practice group to identify sites that were operating at the time of the study.

Data collection followed a rapid qualitative research design involving teams of field researchers, participatory approaches, and iterative data collection and analysis^12 13^. Most of the interviews were carried out by pairs of researchers with different disciplinary backgrounds (i.e. qualitative and quantitative research) to obtain insight into different aspects of the model and their implementation. The interviews were steered by an interview topic guide (Appendix 1), audio recorded and focused on capturing the aims and main components of remote home monitoring models, staff experiences of implementing the models during wave 1 of the pandemic, and processes used to implement the models (including factors that acted as barriers and enablers). The interviews lasted between 30-60 minutes and were carried out by one or a pair of researchers, depending on the interviewee (researchers with qualitative and quantitative expertise sometimes carried out the interview in tandem to capture different types of information).

We obtained information on the data collected during the first wave (including the data fields, numbers of patients covered and aggregate outcomes, and the extent to which the sites used both bespoke and standard data collection). We gathered information on whether they used other available quantitative evidence to help inform clinical decisions. We used this information to assess the value of the data in helping the sites monitor progress against outcomes, understand the impact of virtual wards, and identify the resources used in their implementation.

Documentary analysis was used to capture changes in design and implementation over time as well as to understand key broad contextual factors such as population served, geography and availability of other services.

#### Interview sampling

We identified study sites from early scoping work and conversations with key national leads who were involved in setting up these services in England (combining purposive and convenience sampling). We approached sites that had already established a service and had onboarded patients at the time of the study. We approached eight sites to take part in the study and all accepted this invitation (Table 1). We sought to include sites that functioned as pre-hospital and early discharge from hospital wards, covered different population sizes and areas of the country and had different combinations of monitoring approaches (app, paper-based with telephone calls, and both). In order to be included in the study, the site had to be fully operational during wave 1 of the pandemic.

**Table 1.**
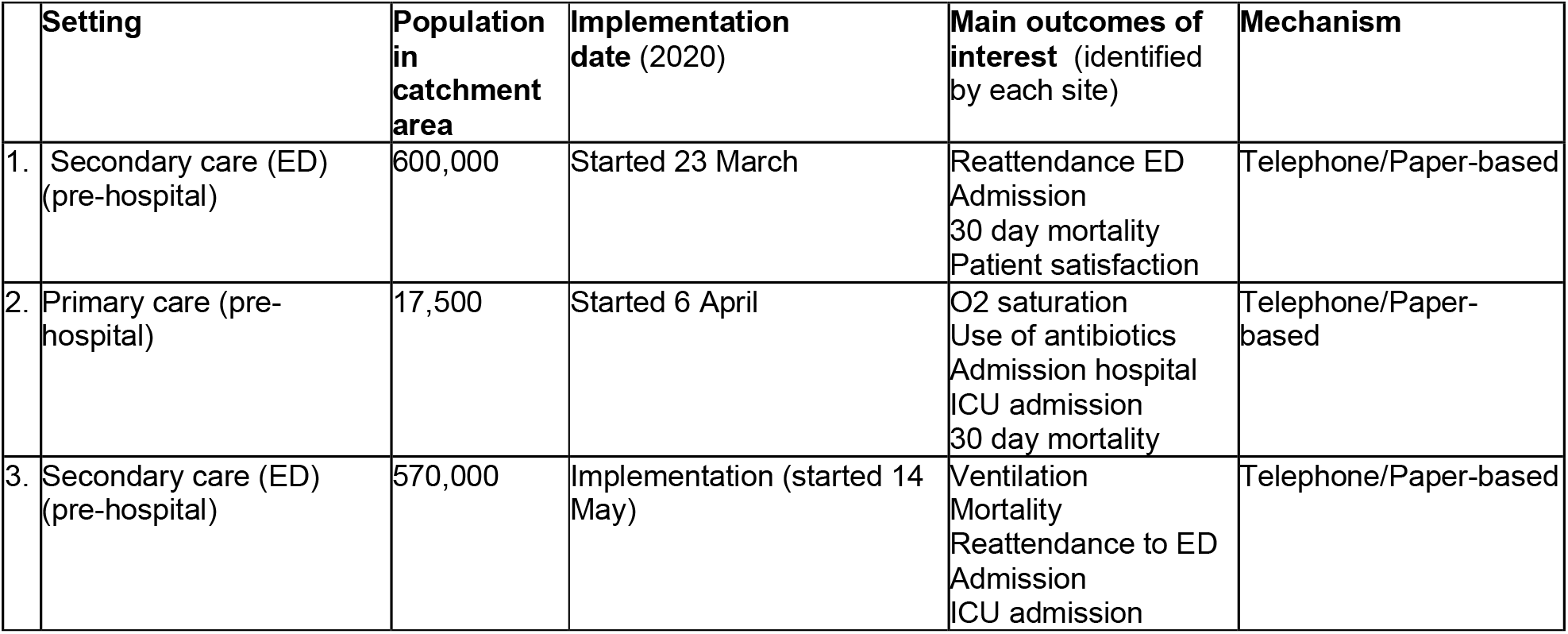
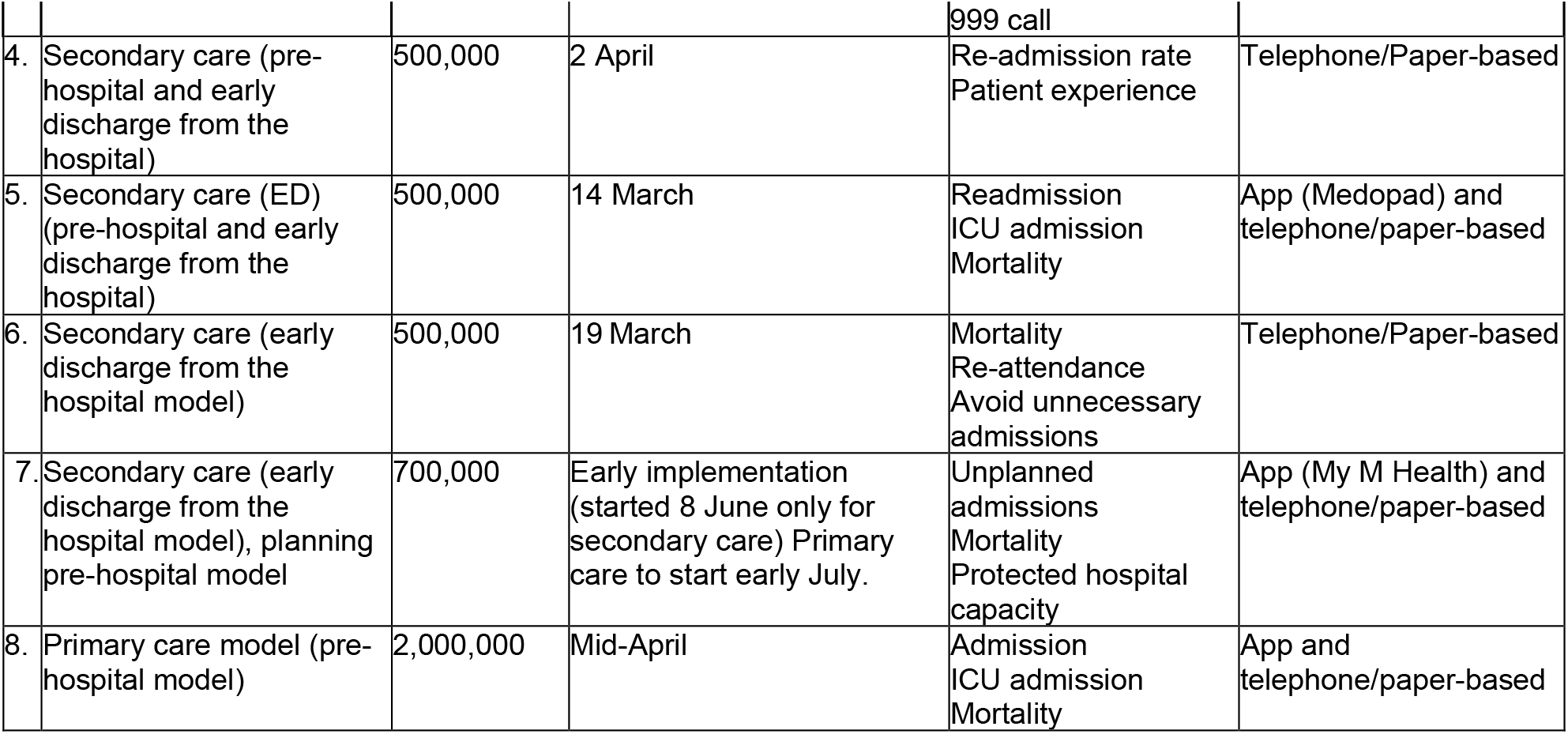
Sample of remote home monitoring pilot sites included in the rapid qualitative study

The interviews were carried out with a purposive sample of study participants based on the flowchart presented in Figure 1. We aimed to carry out interviews with participants who led the design and implementation of the models, staff in charge of monitoring and escalation and staff with knowledge of data collection and use. We approached 25 potential participants and 22 participated in the study, representing a response rate of 88%.

**Figure 1.**
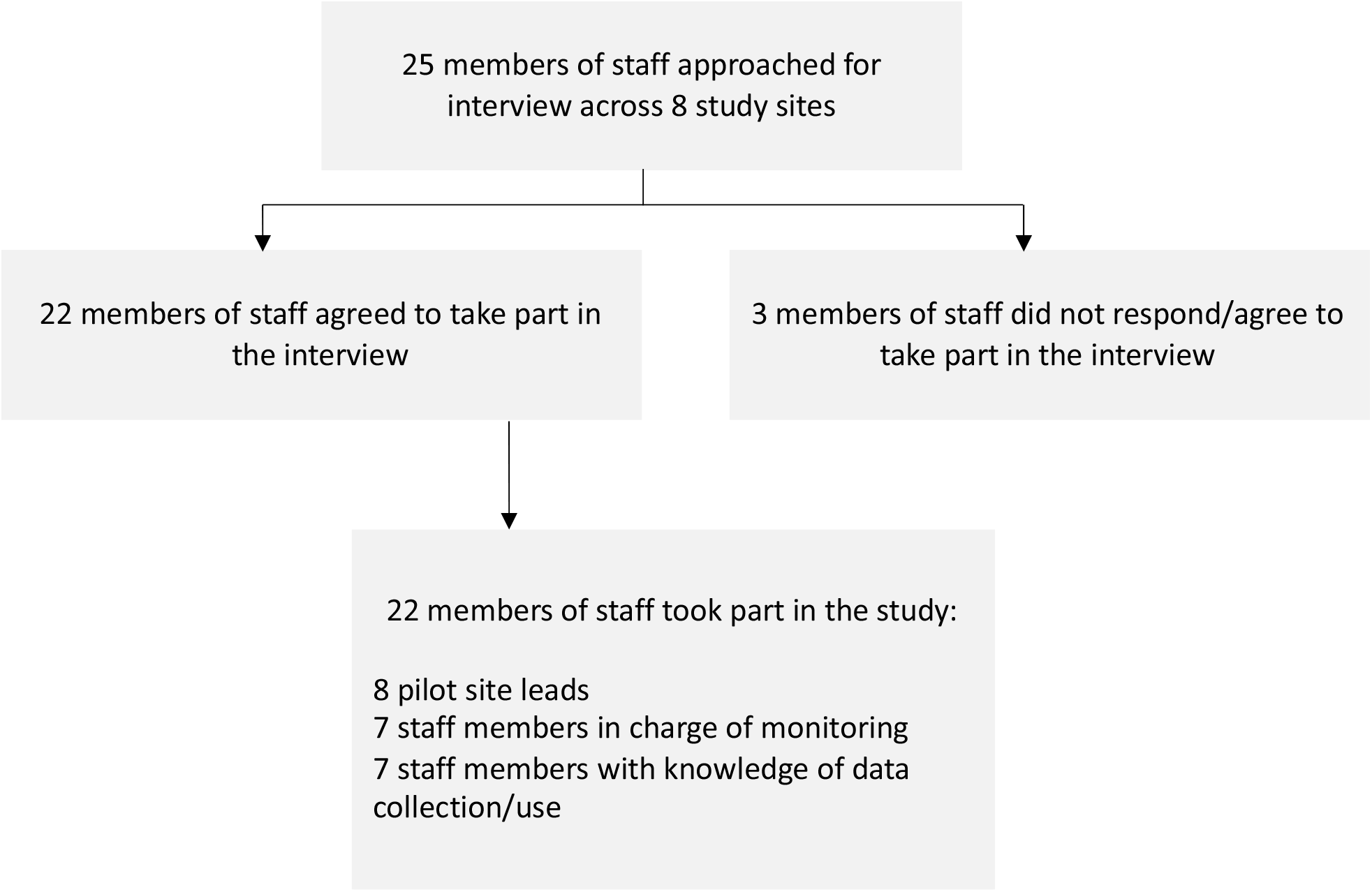
Sampling flowchart for interviews with pilot site participants

#### Recruitment

Site leads guided the researchers in the identification of potential interview participants. The researchers followed an informed consent process by contacting potential participants via email and sending them a participant information sheet (informing them about the study, the anonymity of the collected data and that their participation would be voluntary). Participants were given 48 hours to review the information and ask questions about the study. If the participant agreed to take part in the study, they were asked to sign the consent form. The researcher then arranged a time to carry out the interview over the phone or online. Only members of the team had access to the data.

#### Data analysis

Detailed notes from the telephone/online interviews were added to Rapid Assessment Procedure (RAP) sheets that sought to summarise the main findings from each study site (Appendix 2). We have described their use in detail in Vindrola-Padros et al.^13^. Multiple researchers cross-checked the data included in the RAP sheets to ensure consistency in data reduction and synthesis. The interviews were not transcribed verbatim, however, the team held weekly meetings throughout the study to discuss emerging findings and develop themes based on the data. The team used the RAP sheets to identify the key findings and look for patterns across the study sites. An iterative approach to data analysis was followed, engaging with published literature, the wider NHS Covid community and case study sites for member validation (cross-checking of the interpretation of findings by study participants) and additional insight into the study findings.

### The number of patients and the impact

Seven out of eight sites provided information on the total number of patients served during the period March – August 2020. Using a data collection template (Appendix 3), the total number of patients was collected separately by activitiy (triaged, monitored, deteriorated and escalated). For one site, limited information on the number of patients was obtained from the interviews. In addition, the number of deaths and the number of patients discharged from the virtual wards were also collected.

### Staffing models and resource allocation

We collected data on the staffing models and allocation of resources. This included data on the number of staff, their function/ seniority and the number of hours worked. Data on all other resources used in setting up the pilots and running them were also collected. These included medical equipment used, information materials, tools for flagging deterioration, etc. (Appendix 3). The sites also reported the associated costs to these resources. Six out of eight sites provided the requested information.

#### Data analysis

The information from the pre-hospital and early discharge from the hospital models was analysed separately. The set-up costs were reported as a mean cost per site (as the running period may be longer than the one observed in this study and some of the equipment like oximeters and thermometers can be reused for longer than the study time). For the running costs, the mean costs per patient were calculated for the pilot period (March-August 2020). Here the oncosts like admission to the hospitals, or the ICU costs were not considered because of data limitations.

## ETHICAL APPROVAL STATEMENT

The study was reviewed and classified as a service evaluation by the HRA decision tool and the UCL/UCLH Joint Research Office. It was also reviewed by the University of Birmingham Research Ethics Committee (REC): ERN_13-1085AP37. We followed an informed consent process with patients and obtained written consent before the interview.

## RESULTS

### The design of remote home monitoring models

The main aim of these models was to monitor patients considered high-risk who could be safely managed at home to: 1) avoid unnecessary hospital admissions (appropriate care at the appropriate place), and 2) escalate cases of deterioration at an earlier stage. In most cases, staff drew from experiences of implementing previous remote home monitoring or ambulatory care pathways and used staff familiar with these pathways to deliver the home monitoring.

The steps or main components of these models included (Figure 2):

**Figure 2.**
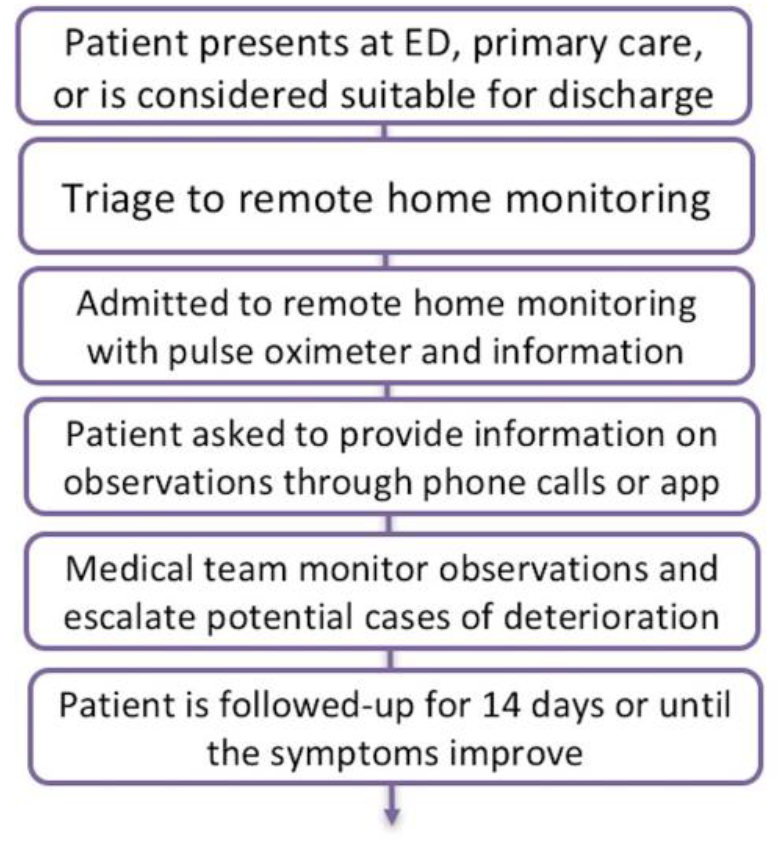
Main steps involved in remote home monitoring models.

1. Assessment of patient for suitability for remote home monitoring. The patient was triaged through emergency telephone numbers, GP practice, (or ED for those pilots in secondary care). Different sites developed their own inclusion criteria. Community-led programmes prioritised higher risk patients (selected by, e.g. age, comorbidities). Hospital-led programmes were concerned with the safety of someone being sent home: oxygen saturation levels and NEWS2 scores were often included as indicators. Some of these models functioned as early discharge from hospital wards in the sense that they were used to monitor patients who had been admitted to hospital but were thought to be fit enough to complete their recovery at home.
2. The patient was provided with a pulse oximeter, patient information (including escalation warning signs and what to do) and a mechanism for recording observations regularly (app or paper diary) such as changes in symptoms, pulse, heart rate, temperature and blood oxygen levels. In some cases, patients were risk-stratified and this informed the frequency of the monitoring.
3. The patient received regular monitoring calls from staff (either primary or secondary care depending on the site) capturing changes in symptoms and trends in oxygen saturation. The modality and frequency of surveillance was based on clinician discretion and calls were used to identify cases of deterioration and inform the patient of next steps. Some sites used apps for patient monitoring, where patient recordings were displayed on a dashboard the clinical teams could access on a continuous basis. The apps had an additional safety mechanism, where these alerted the patient when recorded data indicated potential deterioration and the patient was instructed about next steps (emergency telephone numbers, GP call, etc.). Alerts could also trigger action by the clinical team monitoring patient information on the electronic dashboards, mainly through a phone call to the patient to indicate that they would need to visit a local ED or dial the emergency phone line.
4. Patients were expected to be discharged from remote home monitoring around 14 days (when recovery was expected), but this varied by study site. In some cases, patients were followed-up in person at their local GP practice. Some study sites had linked data to monitor outcomes in patients admitted to hospital.

Three of the models included in the study relied on the use of telephone calls and paper diaries for patient monitoring, while the other three used the telephone/paper diary option as well as an app. Two of the sites using the app used the same app (Medopad/Huma) an app that had been used for the remote monitoring of other conditions prior to COVID-19 and the other sites used the my M-health app, which was still under development at the time of the study.

### Patient experience and engagement

Seven of the eight study sites documented patient experience through surveys or questionnaires. These questionnaires were developed by each site and a national standard or repository for patient experience of remote home monitoring was missing at the time of the study. In general, patient experience was described as positive. Staff described high levels of patient engagement and reports by patients that the service provided reassurance. It is also important to note that staff also reported some cases of increased patient anxiety and the reduction in patient engagement during later follow-up calls or at later stages of the first wave of the pandemic.

### Data and evidence

The study sites collected combinations of demographics, clinical readings, patient experience and outcomes data. Common outcomes collected included hospital and ICU admissions or readmissions, ED attendances, mortality rates and patient satisfaction measures. The need to act quickly at the start of the pandemic meant that there was little time to carefully plan data collection. Study sites aspired to share data between and within primary, secondary and other care sectors, but had made limited progress. Data quality was reported in some sites to be good, while others acknowledged limitations, especially early on. Data collection outside the apps could be cumbersome, and study sites found it challenging to integrate data from apps into their existing patient administration systems. There was relatively little external evidence available on the effectiveness of different approaches that could inform the set-up of the models.

#### Use of data

Some study sites reported that analysis of their own data had helped to inform improvements to their service, and most were monitoring key outcomes over time. One site with a small number of patients acknowledged the difficulty of measuring the impact of their service, and queried whether combining data across different study sites would be possible. No study site had been able to identify an appropriate group to use as a comparator at the time of the study.

More sophisticated analyses of the data were reported to have started with partner health and academic organisations, for example, two sites were collaborating on the development of a prediction model linked to oximetry readings. Further measures that sites would like to have collected included data on the longer-term effects of COVID-19 on patients, and data on the mental health impacts on both staff and patients.

### Patient numbers and impact

The information on the number of patients monitored for the period March-August 2020 for seven out of eight sites (for both the pre-hospital and the early discharge from hospital models) is presented in Table 2. Our complete sample consisted of patients that were monitored remotely. The number of patients triaged was higher than the number of patients monitored. The most plausible reason for this was that some of the patients were not eligible for remote monitoring. However, due to the limitation of quantitative data we had this could not be explored further. The total number of escalated patients was 10% for the pre-hospital and 12.2% for the early discharge from the hospital model, from which the majority of patients was seen in ED (76.7% and 91.8% respectively) and/or admitted to hospital (52.7% and 74.5%). The number of escalated patients admitted to ICU was 2.0% for the pre-hospital and 8.5% for the early discharge from the hospital model. The reported mortality rates (1.1% and 0.9% respectively) were lower than the Case Fatality Rate (CFR – the proportion of people diagnosed with Covid-19 who die) of 6.57% reported in the national data during the first peak^14^. The results are comparable with other similar study in Brooklyn Methodist Hospital where the mortality rate for outpatient telehealth follow-up Covid-19 patients was 1.2%^20^.

**Table 2.** The number of patients

| Throughput and outcome | Pre-hospital Model |  | Early discharge from the hospital Model |  |
| --- | --- | --- | --- | --- |
|  | No. of patients | % of monitored patients | No. of patients | % of monitored patients |
| Patients triaged | 1,861 | 107.1% | 354 | 102.1% |
| Patients remotely monitored | 1,737 | 100.0% | 347 | 100.0% |
| Patients deteriorated and escalated | 174 | 10.0% | 42 | 12.2% |
| Deaths | 20 | 1.1% | 3 | 0.9% |
| Discharged alive from remote monitoring service | 1,639 | 94.4% | 320 | 92.2% |
| **Patient deteriorated and escalated** | **No. of patients** | **% of deteriorated and escalated** | **No. of patients** | **% of deteriorated and escalated** |
| Seen in ED | 133 | 76.7% | 39 | 91.8% |
| Admitted to hospital | 92 | 52.7% | 41 | 74.5% |
| Admitted to ICU | 3 | 2.0% | 4 | 8.5% |
| Treated in primary care | 17 | 17.7% | 17 | 44.7% |

**Source:** Based on data provided by each sites.
**Note:** Data available from 7/8 sites. During this period of time: 5 sites use pre-hospital model only; 1 site early discharge from the hospital model only; 1 site both pre-hospital and early discharge from the hospital models.

### Staffing models and costs

Data on the staff involved in setting-up and running the models did not show clear patterns in terms of specialisation or seniority of the staff involved. The total number of the staff involved in setting up and running the models varied by site. Staff involved were a mix of consultants, ED staff, GP partners, nurses, ANPs, and medical students (Appendix 4 for more information on the staff’s level of seniority and the number of hours spent).

Table 3 gives detailed information on the resources used during the setting-up and running periods for both pre-hospital and early discharge from hospital models. As expected, the set-up period was more resource intensive, while the running was more staff-time intensive. The running costs depended on the time the pilot site was operating and the number of patients seen. The mean costs per monitored patient were higher in the pre-hospital (£553 per patient) than in the early discharge from hospital model (£400/patient).

**Table 3.**
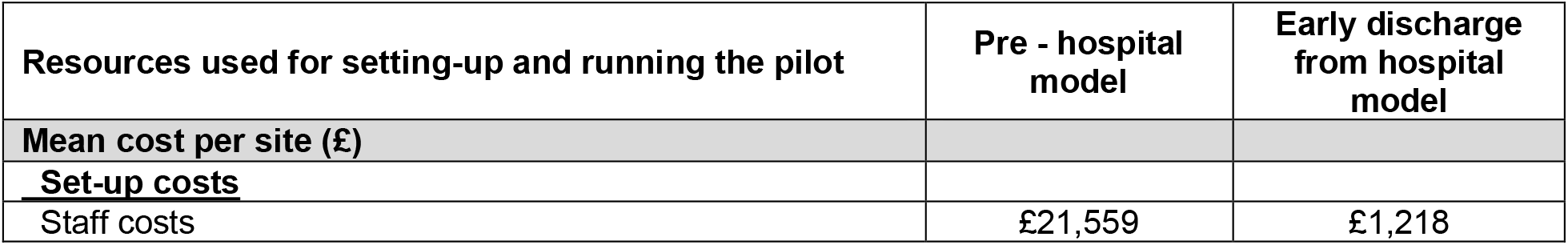
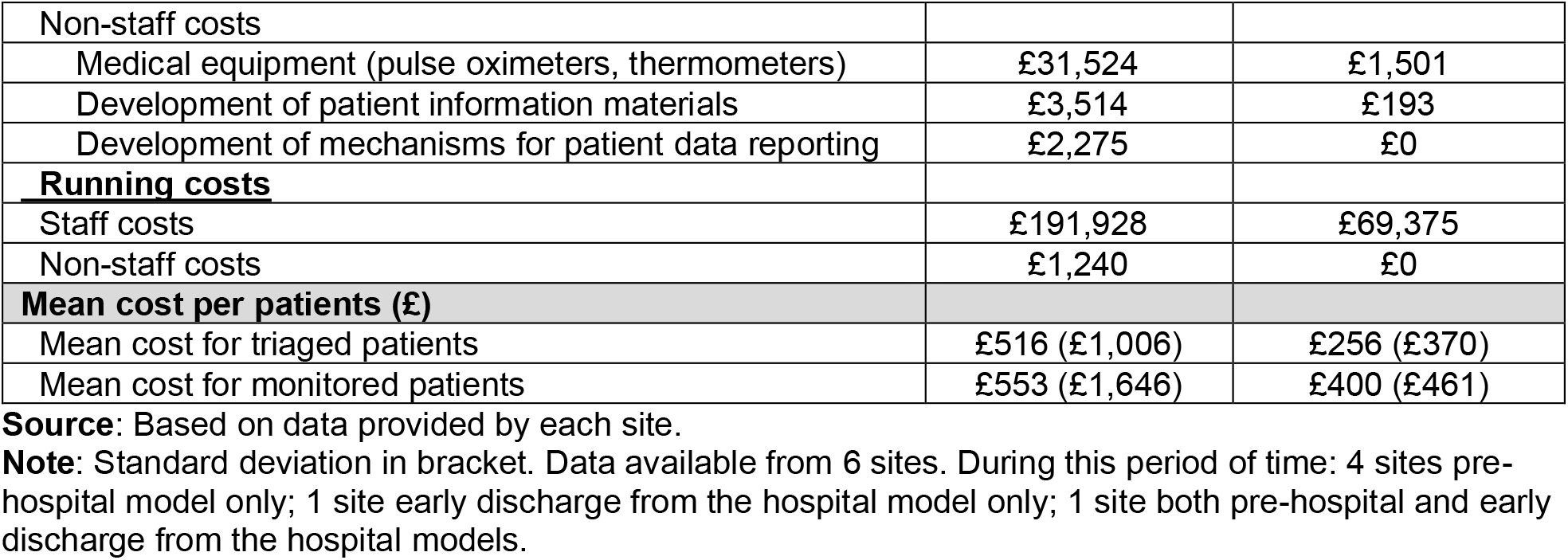
Costs of pre-hospital and early discharge from the hospital models

### Implementation facilitators

Implementation was facilitated by the active role played by dedicated clinical leaders in establishing the remote home monitoring models. Significant support and ‘buy in’ from senior management within acute trusts and across Clinical Comissioning Groups (CCGs) to set up the models was documented across all sites. Acute hospitals that had previous pathways in place (i.e. ambulatory care) or digital protocols that could be repurposed by IT teams were able to set-up these models at a quicker pace. In both primary and secondary care-led models, participants indicated that monitoring could be delivered by nurses with minimal senior oversight, maintaining clear communication with delivery teams. Good communication between members of the clinical team was identified as a key facilitator. It is important to note that during the first wave of the pandemic, staff were available to play a role in the delivery of care in these models due to the cancellation of elective care and other activities in the NHS. Volunteers were also used. Participants expressed concern that these staff members would not be available during future surges in patient cases.

In general, patients experienced positive engagement with the remote home monitoring models. Paper and video patient information (as well as using digital platforms) was very useful to explain the concept of the remote home monitoring models and how to take measurements using pulse oximeters.

### Implementation barriers

Early on, referral criteria and processes were unclear, which led to patients being referred to these models who might have have been ineligible in other circumstances. In part, this was caused by evolving criteria for patient referrals. Staff found it difficult to carry out non-verbal assessments using telephone and video consultation alone. Some patient groups were more difficult to monitor remotely (e.g. homeless community) and staff reflected that monitoring using an app only model might not be suitable for all populations, as this approach could exclude patients with low levels of health and technology literacy. The availability of culturally appropriate patient information in different community languages was identified as a key component of patient engagement, but not all study sites were able to develop these materials.

Lack of administrative/project management support and resources meant that essential equipment such as pulse oximeters could not be obtained quickly. Staff also found it challenging to deliver a seven day service due to workforce availability. There was a lack of published data to support the design of the remote monitoring models and study sites found it challenging and time consuming to collect the desired data, even when using commercially available apps. The integration of service data with existing patient administration systems was generally poor, and it was not feasible to arrange data sharing between and within sectors in the time available. Additionally, there was no link between NHS Test and Trace systems and the study sites’ referral processes.

## DISCUSSION

The monitoring of patients remotely was perceived by staff as a safe way to ensure patients received the appropriate care at the right place. The mortality rate (1%) appeared low, especially when compared to other COVID-19-related mortality rates; but caution needs to be taken when drawing comparisons as populations, pre-existing conditions and levels of severity are likely to be different. Furthermore, these models were not able to establish control groups to compare effectiveness. This is consistent with the published evidence on the use of remote home monitoring models for COVID-19 cases in other countries^11^. Our selection of the models included in the study was based on a combination of purposive and convenience sampling (limited by our knowledge of the models in operation at the time of the study), representing an additional limitation of the study. It is also important to consider that the data generated through the interviews and data collection templates could have been shaped by self-reporting and recall bias. Models of care were decided at haste given the severity of the situation so questions about comparators were not considered a priority.

Patient and carer training were identified as the key to the success of these models and some concerns were expressed in relation to models that only relied on digital solutions such as apps for the monitoring of patients and the identification of cases of deterioration. Technological barriers have been reported in studies of remote home monitoring for other conditions and authors have highlighted the impact of these barriers on patient engagement and outcomes^15 16^.

The development of inclusive models was discussed frequently, and, to some extent, this was reliant on the availability of patient information that was culturally appropriate and in different languages. Some published examples of the use of pulse oximetry in remote home monitoring models have indicated that good patient training is associated with more accurate readings and more responsive care^5^. Personalised support might be required to avoid patient anxiety and reach those who may be difficult to monitor remotely. Remote home monitoring models in the US and Canada, for instance, also screened patients for mental health and social care needs, providing a more holistic model of patient care^4 17^.

Our study also found concerns regarding the sustainability of services as the models we analysed were implemented during the first wave of the pandemic, when some members of staff were released from clinical responsibilities and could be redeployed to monitor patients remotely. Moreover, our study did not explore the extent to which remote home monitoring created additional workload for the staff involved, flagged previously by another study conducted in the Netherlands.^18^. Furthermore, additional sources of funding were made available at this time and staff were allowed to use discretionary funds through fast-approval processes established in both primary and secondary care. The sustainability of these models during subsequent surges in patient cases and for other conditions will require more stable flows of funding as well as clinical and administrative management support. The detailed information on the use of resources was limited and more resources could have been used for set-up and running of the services, but not reported (including the costs of admission to the hospital or the costs of ICU). It was also difficult to conclude if the differences in resource use between the sites were a result of barriers/availability of resources or reflected the specific patterns of use in these different contexts.

The impact of remote home monitoring on patient outcomes and their cost-effectiveness should also be assessed through the use of more standardised data and appropriate comparators. Future research will also need to explore the benefits and limitations of models led by primary versus secondary care to determine if primary care is able to deliver more integrated care for patients and reduce the demand on hospital services. The analysis of patients’ and carers’ experiences with these models should be a core component of future studies, and may shed further light on how best to engage and support patients to maximise the effectiveness of remote home monitoring^18^. As the healthcare landscape continues to adapt to the challenges generated by COVID-19 and the NHS enters different seasonal peaks, future studies will need to analyse the transformation of these models over time^19^, beyond the pressures and resources made available during the first wave of the pandemic and considering changes implemented in the risk stratification of patients and Test and Trace programmes.

## Data Availability

All data relevant to the study are included in the article or uploaded as supplementary information.

## CONTRIBUTORSHIP STATEMENT

NJF, CSJ, TG, KS, MS, SMT, SM and CVP contributed to the design of the study. MS, KS, CSJ, TG, SMT, NJF and CVP participated in data collection for the study. NJF, JE, and SM reviewed and provided feedback on the manuscript. All authors approved the final version of the manuscript. The corresponding author attests that all listed authors meet authorship criteria and that no others meeting the criteria have been omitted.

## ACKNOWLEDGEMENTS

We would like to thank our clinical advisory group, Dr Matt Inada-Kim, Dr Allison Streetly and Dr Karen Kirkham, for their advice and support throughout the duration of the study. We would also like to thank all of the staff members who gave of their time to participate in the study.

## Appendix 1. Interview topic guide

### A rapid service evaluation of remote home monitoring (‘virtual ward’) during the COVID-19 pandemic in the UK

1. Can we start with a description of your current role?
2. How did the remote home monitoring model or pilot originate and how did it develop over time?
  a. *Why were remote home monitoring pilots needed at the time?*
  b. *Did it develop from a previous remote home monitoring model or ambulatory care pathway?*
  c. *How long did it take you to get the model up and running?*
  d. *Who led the development of the model?*
3. What were the aims of the model and its main features?
  a. *Population served*
  b. *Characteristics of patient groups*
  c. *Pre-hospital, step-down or both?*
  d. *Availability of other services at community or primary care level*
4. What were the processes involved in the pilot?
  a. *Patient triage*
  b. *Patient information and training*
  c. *Patient monitoring (what was monitored and how)*
  d. *Mechanisms used for patient data reporting (i*.*e. app, paper-based)*
  e. *Tools for flagging deterioration*
  f. *Escalation processes*
  g. *Patient discharge from ward*
5. What were the outcomes you expected from the implementation of the pilot? How did you think the pilot would help you achieve these outcomes?
6. What were the factors that acted as barriers and facilitators in the design and implementation of the pilots during wave 1 of the pandemic?
7. What data did you collect on patients while implementing the pilot?
  a. *Have these data been linked to other data sources?*
  b. *Who is able to access these data?*
  c. *How have these data helped you to monitor progress against your expected outcomes?*
  d. *Have there been changes to the data you’ve collected during the pilot? If yes, why were changes made?*
  e. *Would you be able to share aggregated data (i*.*e. total number of patients, their outcomes, etc*.*), and lists of data fields with us?*
8. What data or information, if any, would you have liked to have collected, but couldn’t? Why?
9. What quantitative evidence, if any, did you make use of to design parts of the pilot processes and inform your decisions?
10. What resources were allocated (including staff) to implement the pilots? Do you have any documents you can share with us?
11. What was the staffing model used for the pilot?
  a. *Staff grades*
  b. *Rota*
  c. *Responsibilities*
12. What is your perception of patients’ and carers’ experiences of the pilot?
  a. *Did you have trouble accessing any patient groups?*
  b. *Did you feel any patient groups had difficulties accessing care or being onboarded onto the virtual ward?*
  c. *Do you feel patients and carers received all of the necessary information? Do you feel they understood the information?*
  d. *Were patients able to engage well with the services delivered throughout the pilot?*
  e. *Did you collect any data on patient experience? If so, are you able to share this with us?*
13. What are the lessons learnt from implementing the pilot during wave 1 of the pandemic?
  a. *Benefits of these models and areas of good practice*
  b. *Limitations of these models*
  c. *Sustainability of the models*
  d. *Areas that need to be improved*
  e. *Addressing inequalities in patient access*
14. Did you adapt the model after wave 1 of the pandemic or do you plan to adapt it now? If so, why and in what ways?
15. Would you do anything differently during wave 2?
  a. *Patient triage*
  b. *Patient information and training*
  c. *Patient monitoring*
  d. *Mechanisms used for patient data reporting (i*.*e. app, paper-based)*
  e. *Tools for flagging deterioration*
  f. *Escalation processes*
  g. *Patient discharge from ward*
  h. *Staffing model*
  i. *Guaranteeing patient access and addressing inequalities*
16. What advice would you give colleagues attempting to implement similar pilots in other areas of the country?
17. Is there anything else you think we should know that I have not asked you?
18. Would you be interested in participating in phase 2 of the evaluation?

## Appendix 2. RAP sheet

| Topic | Main findings |
| --- | --- |
| General context |  |
| Development of the pilot (history of why it was developed and how it changed) |  |
| Aims and main features of the pilot |  |
| Components/processes involved in pilot |  |
| Target patient population/s |  |
| Implementation stage |  |
| Barriers and facilitators during implementation |  |
| Data<br>(data collected, linkage of data, expected outcomes, use of data, who has access to data, if data has changed, data important but did not collect, able to share aggregated data) |  |
| Resources allocated<br>(funding, staffing, etc.) |  |
| Staffing model<br>(Staff grades, rota, responsibilities) |  |
| Perception of patients' and carers' experiences<br>(Trouble accessing patient groups, information for patients, patient engagement, data on patient experience) |  |
| Lessons learnt<br>(Benefits, limitations, sustainability areas that need to be improved) |  |
| Current state of pilot |  |
| Adapting model after wave 1 |  |
| Things would do differently in wave 2<br>(Triage, patient information and training, monitoring, reporting, flagging deterioration, escalation, discharge, staffing model) |  |
| Advice to others |  |
| Potential evaluation questions |  |
| Interested in taking part in phase 2 |  |
| Other comments/reflections |  |
| Number of patients served for the period April 2020 – 31 August 2020 |  |
| The number of staff involved in setting up the pilot for the period April 2020 – 31 August 2020. |  |
| The resources used for setting up the pilot for the period April 2020 – 31 August 2020 |  |
| Implementing the Pilot: The job band of the members of staff undertaking each of the activities |  |
| Other comments: |  |

## Appendix 3. Data collection template

### Evaluation of implementation of RM pilot virtual wards for COVID-19 patients: data collection form for resources, staffing, and impact

#### A) Patient numbers and outcomes

1. Please complete the numbers in table below.

| Activities | Total number of patients during the period covered |
| --- | --- |
| I. Patient triaged |  |
| II. Patient monitored |  |
| III. Patients deteriorated and escalated |  |
| How many of the patients from the latter group (deteriorated and escalated) were advised to: |  |
| (i). Dial 111 |  |
| (ii). Dial 999 |  |
| (iii). Present to ED |  |
| How many of the deteriorated and escalated patients were: |  |
| (i). Seen in ED |  |
| (ii). Admitted to hospital |  |
| (iii). Admitted to ICU |  |
| (iv). Treated in primary care |  |
| IV. Number of deaths |  |
| V. Number discharged from the virtual ward |  |

#### B) Setting Up the Pilot

1. What type of staff were involved in **<u>setting up the pilot</u>**, and approximately how much time did they spent?

B) Setting Up the Pilot 1. What type of staff were involved in setting up the pilot, and approximately how much time did they spend?
| Staff involved in <u>setting up the pilot</u> | Staff's band/function | Number of staff members involved | Number of total hours spent per staff |
| --- | --- | --- | --- |
| (i). |  |  |  |
| (ii). |  |  |  |
| (iii). |  |  |  |
| (iv). |  |  |  |

2. What other resources were used in **<u>setting up the pilot</u>**, and approximately how much did these costs?

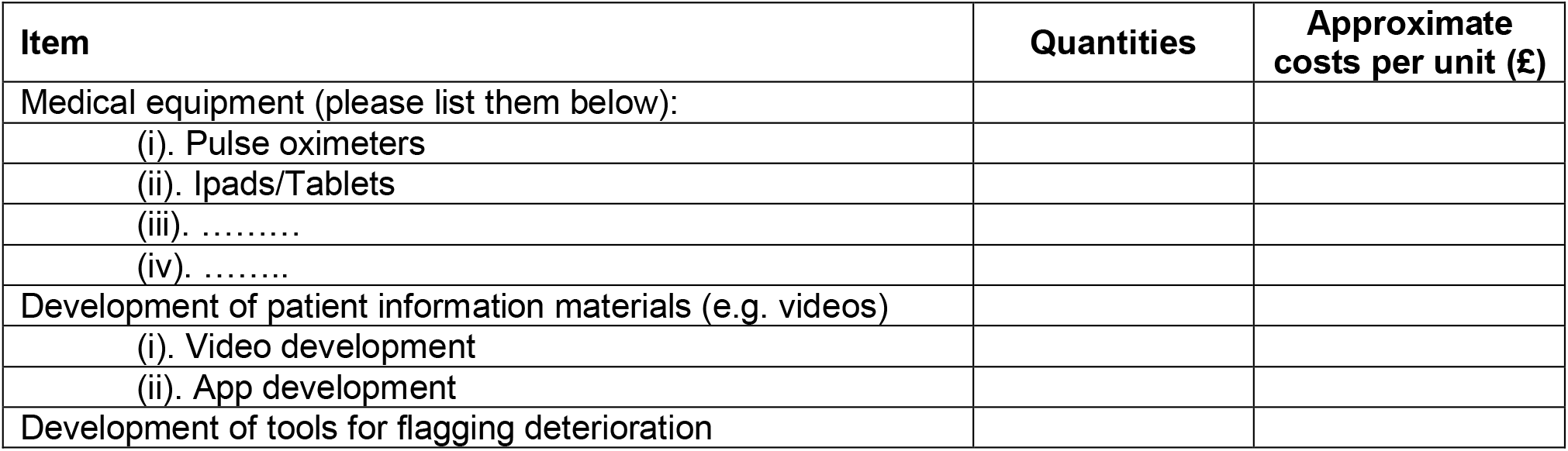
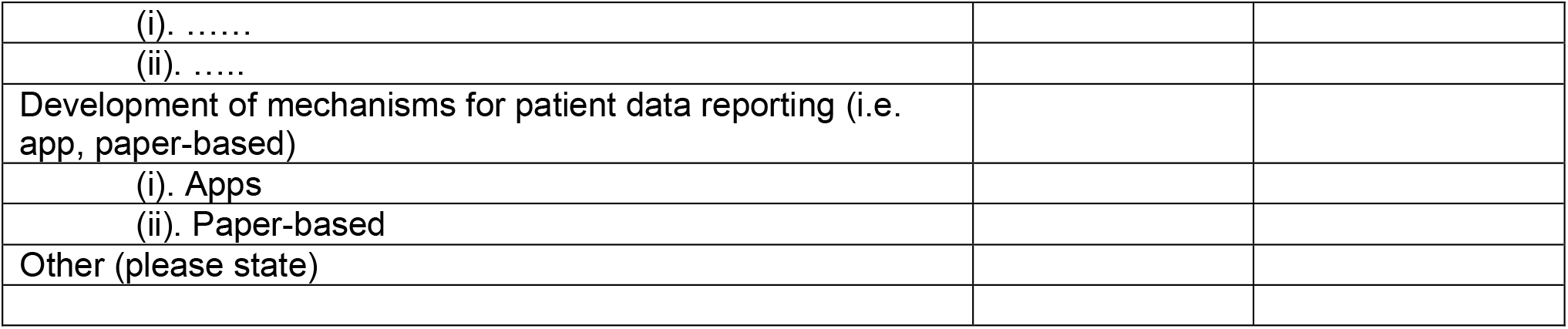

#### C) Implementing the Pilot

1. What is the job band of the members of staff undertaking each of the activities below and how many hours/shifts did they spent on average per week.

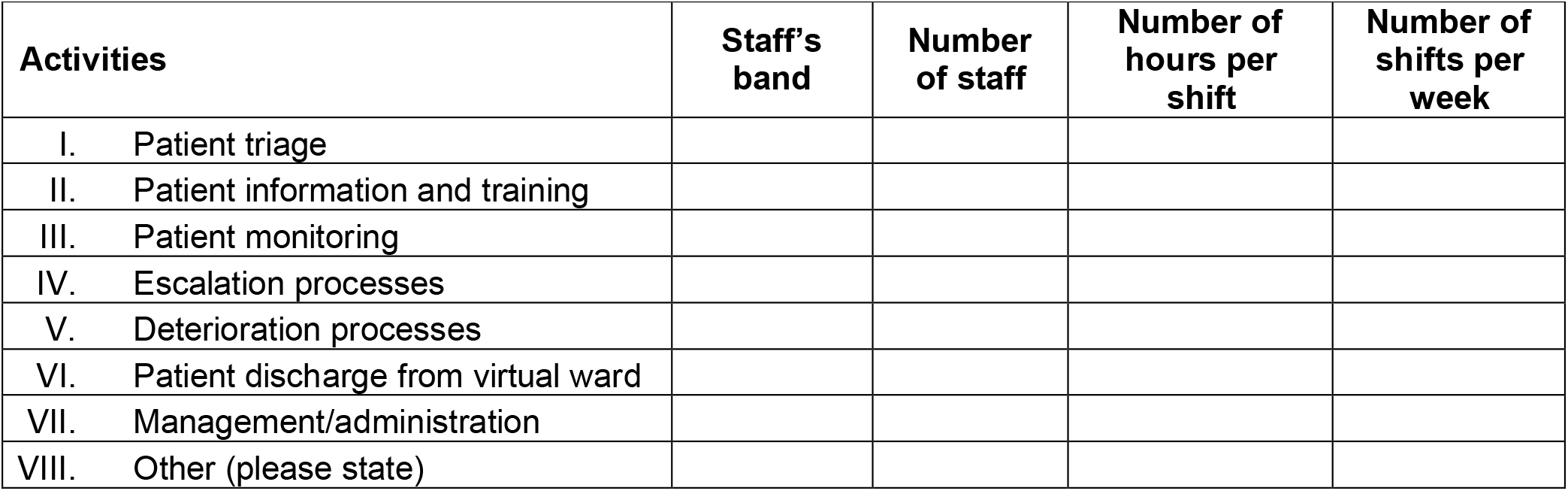

2. Other than staff time, are there any other resources (e.g. equipment) used for each of these activities during the period covered. If so, please record this below:

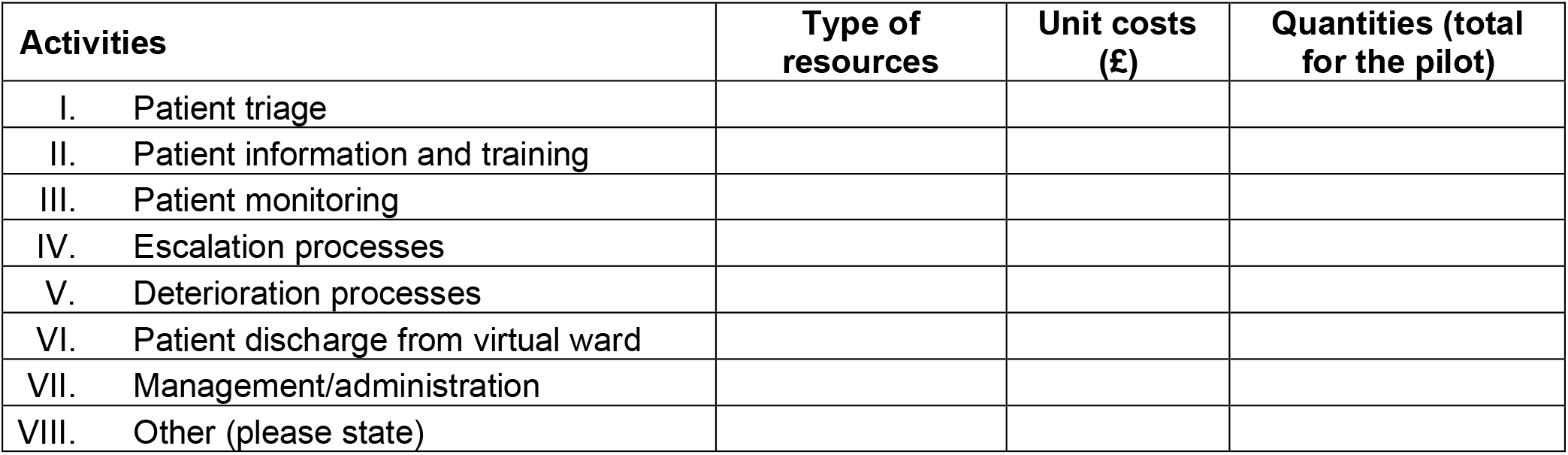

## Appendix 4. Staffing models

**Table 1.**
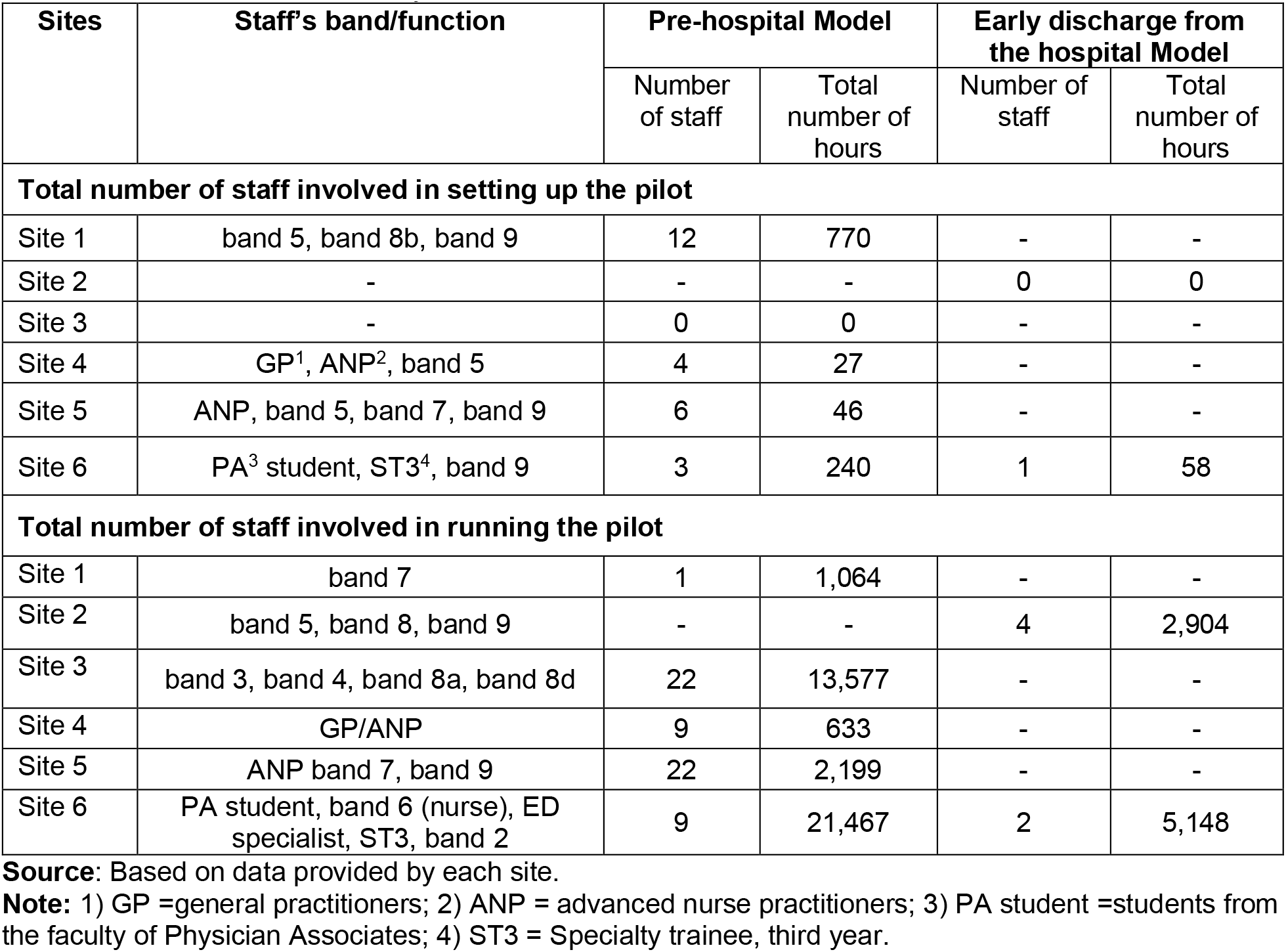
Resource allocation by site

**Table 2.** Staffing models by site

| Sites | Setting |
| --- | --- |
| Site 1. | Secondary care: patients monitored remotely by respiratory nurses<br>Primary care: delivered using federation funded primary care nurses who have worked in the community |
| Site 2. | Specialist consultant, a band 8 nurse, Outpatient Parenteral Antibiotic Therapy (OPAT) nurses who were redeployed helped with the calls (during peak also relied on help from specialist respiratory nurses). |
| Site 3. | A mix of consultants, cardiologists, five physiotherapists, three physiologists, house officers (largely data collection and completing telephone calls); medical secretaries, and a medical student setting up and monitoring the database |
| Site 4. | Small core team of GP partners, practice manager and ANP; led by senior GP partner |
| Site 5. | Delivered using three ANPs, senior and junior clinicians, and specialist registrar conducting data collection and analysis. |
| Site 6 | One ICU consultant, 1 ultrasound fellow, 1 medical student, 4 PAs, admin support and 3 furloughed middle grade ED doctors (from high-risk groups) provided assistance |
| Site 7. | Pilot lead, staff in the ED (they were able to get involved in the pathway as some of their normal work had been paused), and an ED registrar was involved in phoning patients. |
| Site 8. | Pilot lead and model delivered mainly by nursing staff based in primary care. |

